# Factors that influence-COVID-19 Vaccine Uptake and Hesitancy Among a Population in the West Department of Haiti: Implications for Enhancing Effectiveness of Immunization Programs

**DOI:** 10.1101/2024.01.10.24301096

**Authors:** Martine Etienne-Mesubi, Babatunji Oni, Nancy Rachel Labbe-Coq, Marie Colette Alcide-Jean-Pierre, Delva Lamarre, Darwin Dorestan, Marie-Ange Bien-Aime, Venice Dorce, Cory Freivald, Cowan Angell, Yingjie Wang, Jenevieve Opoku, Bryan Shaw, Deus Bazira

## Abstract

**Introduction:** The COVID-19 pandemic in Haiti led to increased challenges for a population concurrently dealing with natural and social disasters, poor quality health care, lack of clean running water, and inadequate housing. While half a million vaccines for COVID-19 were donated by the United States to the government of Haiti, less than 5% of the population agreed to be vaccinated. This resulted in thousands of unused doses that were diverted to other countries. The purpose of this study was to evaluate population characteristics related to vaccine uptake in order to inform future interventions to improve COVID-19 vaccine uptake as well as inform strategies to safeguard against future global health security threats.

**Methods:** This was a mixed-methods, cross-sectional study conducted in the West Department of Haiti within peri-urban communes. Survey participants consisted of adults of this setting responding to an electronic survey between June – Sept 2022. The survey assessed demographic information, household characteristics, religious beliefs, past vaccine use, and current COVID-19 vaccine status. Multivariate regression modeling was conducted to assess predictors of vaccine hesitancy. Qualitative focus group discussions were conducted among community leaders and health professionals to provide additional, community-level context on perceptions of the COVID-19 pandemic and vaccines.

**Results:** A total of 1,923 respondents completed the survey; of which a majority were male (52.7%), were between the age of 18-35 (58.5%), had a medical visit with the last year (63.0%) and received the COVID-19 vaccine (46.1%). Compared to those who had been COVID-19 vaccinated, participants who had not been vaccinated were more likely to be male (57.7% vs 46.8%, p<.0001), have classical education (30.5% vs 16.6%, p<.001), unemployed (20.3% vs 7.3%, p<.0001) and had a medical visit 3 or more years ago (30.2% vs 11.2%, p<.0001). Unvaccinated COVID-19 participants were also more likely to have never received any other vaccine (36.1% vs22.5%, p<.0001), have a religious leader speak out against the vaccine (20.0% vs 13.1%, p<.0001), not believe in the effectiveness of the vaccine (51.2% vs 9.1%, p<.0001) and did not trust the healthcare worker administering the vaccine (35.2% vs 3.8%, p<0.0001).

**Conclusion:** These results show that targeted interventions to religious leaders and health care workers on how to engage with the community and share clearer messages around the COVID-19 vaccination may result in increased vaccine uptake. Results also shed light on how activities surrounding vaccinations can be tailored to meet client needs addressing the misinformation encountered to achieve greater health impact thereby safeguarding the population against future global health security threats.

## Introduction

While the development and implementation of vaccination programs have been a seminal achievement for public health globally, the perception of vaccines as harmful or unneccesary resulting in a hesistancy to take any specific vaccine, has grown overtime (Cooper, 2018; Dube 2013; Sato, 2018). Within the field, the very definition of vaccine hesistancy has evolved over time. MacDonald and colleagues defined vaccine hesitancy as a “delay in acceptance or refusal of vaccination despite availability of vaccination services,” often due to specific regional and cultural contexts (MacDonald, 2015), while others thought this should be defined as a “process of decision-making” (Peretti-Watel, 2015). The dynamics of this phenomenon led to the development of various working groups that targeted vaccine hesitancy and confidence, vaccine demand, and behaviorial and social drivers of vaccinations (NVAC, 2015; Schuster, 2015; WHO, 2014; WHO, 2019). In 2019, the World Health Organization (WHO) cited vaccine hesitancy as one of the top ten threats to global health (WHO, 2019). Various vaccinations such as routine childhood vaccines, HPV, and more recently COVID-19, have improved overall population health. However, these and other vaccines continue to lag in uptake and acceptance across many communities globally (Atwell, 2014; Nguyen, 2022).

Although vaccine hesistancy did not begin with emergence of COVID-19, it has been exacerbated by this threat and continues to be an area of great public health concern. A recent report published by the United Nations Children’s Fund (UNICEF) found that confidence in vaccines for children is currently lower than prior to the COVID-19 pandemic in many countries (UNICEF, 2023). There has been a decline in confidence in childhood vaccines of up to 44% in some countries. This is resulting in a reduction in the protection against preventable diseases such as measles and poliovirus. The WHO has developed the Immunization Agenda 2030, which aspires to achieve 90% global coverage for routine childhood vaccinations while also moving quickly to introduce a new round of immunizations in low-income countries. However, this agenda could be derailed if the factors that contribute to hesitancy, low confidence in vaccine importance, or transparency are not addressed. The erosion of confidence in health care workers, misinformation spreading over social media, as well as other social and cultural factors should be identified and appropriately addressed if we are to meet the goals of, not only vaccinating populations against COVID-19, but also against other preventable diseases.

Haiti has had access to the COVID-19 vaccine since August 2020, however only 3% of the population has been vaccinated for COVID-19 (CARPHA, 2022). The Haitian health system is extremely limited and managing COVID-19 has proven to be an enormous strain on the health care resources. With less than five health institutions in the entire country able to care for patients with COVID-19, a shortage of personal protective equipment (PPE), and an increase in rampant misinformation about the disease, the response has been strained. Additionally, the insecurity challenges that have increased over time have intensified the poor response to health seeking behaviors. The goal of this study was to investigate COVID-19 vaccine hesitancy and vaccine acceptance in the West department of Haiti. Findings will support future efforts in increasing COVID-19 vaccination uptake as well as general childhood and adult immunization efforts.

## Methods

### Study Design and Population

This was a mixed-methods, cross-sectional study conducted in the West Department of Haiti within peri-urban communes. Recuitment for this study started on June 6, 2022 and ended on September 1, 2022. Quantitative data was collected through a survey of adults resident in this setting designed by Georgetown University Center for Global Health Practice and Impact (CGHPI) Haiti Study team. The survey was administered electronically and was used to assess COVID-19 vaccine hesitancy, uptake, and associated factors. Focus group discussions (FGDs) were conducted among community leaders and health professionals to provide qualitative data on vaccine acceptability and hesitancy at the community level. Verbal consent was obtained from all participants prior to study inclusion. Participants were provided with a detailed explanation of the purpose of the study as well as the risks and benefits to study participation. Participants were given an opportunity to ask questions about the study. Verbal consent was documented by the research team. A consent form detailing the study details was also provided to participants for their reference.

### Survey Development, Sampling, Data Collection, Analysis

The survey was adapted from the COVID-19 vaccination acceptability study (COVAccS) (Sherman et al., 2021). Study participants were asked about sociodemographic characteristics such as age, gender, marital status, level of education, religion, employment status, department of residence, and number of persons living in their household. Characteristics describing participants’ health and history of illness were also asked such as the last time that they had a medical consultation, diagnosed with any chronic illnesses, or whether they had ever been vaccinated against any disease. Knowledge of Covid-19 vaccines was also asked, as well as vaccine information sources, including whether participants were aware of the different types of COVID-19 vaccines that were available in Haiti, whether they had been vaccinated against COVID-19, and reasons why they had or had not been vaccinated. Questions about perceived vaccine effectiveness and perceived health worker knowledge about vaccines were also evaluated. The survey also assessed influences of religion and culture on whether one decided for or against getting vaccinated. Similarly, questions about the influence that policy makers or political leaders have on decisions to get vaccinated or to abstain from vaccinations were also assessed.

Eligibility criteria to participate in the study included being a resident within any commune of the West Department of Haiti and being 18 years of age or older. Recruitment of participants was done through administrators of institutions such as churches, voodoo sectors, and health institutions. This was a convenience sample of participants within these sectors. The total target sample size required was 1,583 study participants considering a confidence interval of 95% with a conservative estimate of 50% of Haitians willing to be vaccinated and an expected response rate of 80%. Community health care workers (CHCWs) were trained over a period of two weeks and the survey data collection was completed between June 15, 2022 and July 18, 2022. Participants provided their consent prior to administration of the survey. Surveys were developed on the Qualtrics™ platform and uploaded onto tablets that were used by CHCWs to administer the surveys to participants. Surveys were available to participants in Creole and French and took 30 minutes on average to complete.

Chi-squared tests were used to compare categorical variables and t-tests to compare means between vaccinated and unvaccinated persons. Univariate logistic regression analyses were conducted to assess factors associated with vaccine hesitancy. Our study defines vaccine hesitancy as a delay in acceptance or refusal of vaccination in spite of availability of vaccination services (Macdonald, 2015). Factors found to be significantly associated with vaccine hesitancy were included in the mulitivariate model, adjusting for sociodemographic variables. We present adjusted odds ratios with 95% confidence intervals. SAS v9.2 was used to analyze the survey data.

Qualitative data were collected through 16 FGDs conducted with purposively selected community and religious leaders and health professionals. FGDs were stratified into those that had reportedly received at least one dose of a COVID-19 vaccine and those that refused. Participants discussed their knowledge and perceptions of COVID-19 and vaccines as well as their reasons for/against getting vaccinated. Discussions involved 8-12 individuals, were facilitated by a two trained data collectors in Creole and French, and took 45 minutes on average to complete. FGDs were audio-recorded and translated and transcribed into English. We employed thematic content analysis using the software MAXQDA 2022 (VERBI Software, 2021) for data analysis.

### Ethics Statement

This study was approved by the Georgetown University Institutional Review Board, study number STUDY00004690. The study was also approved by the National Committee of Bioethics in Haiti, Reference number 2122-39. Verbal consent was obtained from all participants prior to study inclusion.

## Results

Of the 1,932 individuals approached to participate in this study, a total of 1923 (99.5%) respondents provided consent and completed the survey. A majority were male (52.7%), were between the age of 18-35 (58.5%) and were married or in a common-law relationship (43.6%). A majority of participants (86%) cited a specific religious affiliation, 96.2% reported some level of education, 65.1% reported some level of employment and 52.3% were living with five or more people in their household.

Compared to those who had been COVID-19 vaccinated, participants who had not been vaccinated were more likely to be male (57.7% vs 46.8%, p<.0001), have classical education (30.5% vs 16.6%, p<.001), unemployed (20.3% vs 7.3%, p<.0001) and had a medical visit 3 or more years ago (30.2% vs 11.2%, p<.0001). Unvaccinated participants were also more likely to be between the ages of 18-25 (32.5% vs 15.8%, p<.0001) and were more likely to be single (57.3% vs 40.1%, p<.0001).

Within our population, we found four main buckets that contributed to certain facilitiators and barriers to vaccine uptake. These include: health seeking behaviors, knowledge of COVID-19 and sources of information, trust in the healthcare system.

### Health Seeking Behaviors

Health seeking behaviors have been associated with various factors such as health literacy, worsened health outcomes or long distance to care facilities (Lubetkin, et. al, 2015, Musoke, et. al, 2014). Our study found that unvaccinated participants were more likely to report not having a medical visit for 3 or more years (30.2% vs 11.2%, p<.0001) and more likely to never have taken a vaccine previously (36.1% vs 22.5%, p<.0001). However, we saw no difference among participants who reported having comorbidities.

About 9.0% of those who were unvaccinated reported distance (9.5% vs. 1.7%, p<0.001) and wait time (9.0% vs. 1.7%, p<0.0001) as being a prohibitive factor to getting vaccinated compared to almost 2% of those where reported receiving the vaccine.

Focus group discussions around this theme supported quantitative findings, revealing that many unvaccinated participants were hesitant to believe COVID-19 existed in Haiti, shared common misconceptions or misinformation around COVID-19 or the COVID vaccine, and/or experienced mistrust in government and health officials. In addition, many participants, vaccinated and unvaccinated, shared concerns over the effectiveness of the COVID vaccine or expressed fear over negative side effects. These factors supported a majority of participants’ health-seeking behaviors or reasons why they had not been vaccinated. Several participants across varying groups of vaccinated and unvaccinated participants shared similar remarks around disbelief in COVID or its existence in Haiti.

> *“I myself do not believe that it [COVID-19] exists for us Haitians”*

Among participants who had been vaccinated, common motivations for seeking a vaccine were protection of self, as required by work or travel, for incentives such as food or money, and/or in response to attending a training on COVID-19 and the new vaccine.

### Knowledge of COVID-19 and Sources of Information

Knowledge of COVID-19 disease and having reliable sources of information may have also played an important role in vaccine hesitancy within this population. Our findings showed that unvaccinated participants were less likely to be aware of the COVID-19 vaccine in Haiti (71.3% vs 19.1%, p<.0001), less likely to believe in the effectiveness of the vaccine (8.1% vs 64.4%, p<.0001) but more likely to have a religious leader speak out against the vaccine (20.0% vs 13.1%, p<.0001). Vaccinated participants were more likely to believe in their source of COVID-19 information (50.2% vs 20.4%, p<.0001). Vaccinated participants also reported that their leaders were more likely to encourage them to take the vaccine (48.8%) compared to those who were not vaccinated (37.6%) p<.0001).

The knowledge of COVID-19 and sources of information among focus group participants in this study reveal a complex landscape of beliefs and concerns. Notably, the analysis on the focus group discussions aligns with our quantitative finding that individuals in vaccinated groups tend to exhibit greater trust in their COVID-19 information sources and beliefs in vaccine effectiveness. However, while they acknowledge the significance of vaccines in safeguarding the community, concerns persist regarding vaccine side effects and myths related to the vaccine. Additionally, there is a prevailing sense of mistrust in both domestic and international authorities, contributing to vaccine skepticism. This skepticism encompasses worries related to unknown side effects, vaccine efficacy, suspicions of political agendas, and the spread of misinformation. This emphasizes the need to work on strengthening government credibility as a go to source for public information. This will enable accurate information dissemination and equitable vaccine access to combat misinformation and promote vaccinations in Haiti. One respondent expressed concern about the situation:

> *“ … we know that we live in a country where the state does not exist - already during the cholera epidemic, we saw that the health actors and the state’s response is still very very weak, not what it should be. So my concern is that our state will not react as it should react, then the population will not be reactive enough to report the damage the disease can cause and the behavior of the population could favor the disease and spread it even more..”*

### Trust in the Healthcare System

Trust in the healthcare system and manufacturing industries were also evaluated among Haitian residents in relation to vaccine hesitancy. Unvaccinated participants were more likely to distrust the pharmaceutical industry (65.4% vs 25.0%, p<.0001), distrust that the vaccine is good for their health (47.3% vs 12.7%, p<.0001) and more likely to distrust healthcare workers who administer the vaccines (35.2% vs 3.8%, p<0.001).Table 1.

**Table 1.**
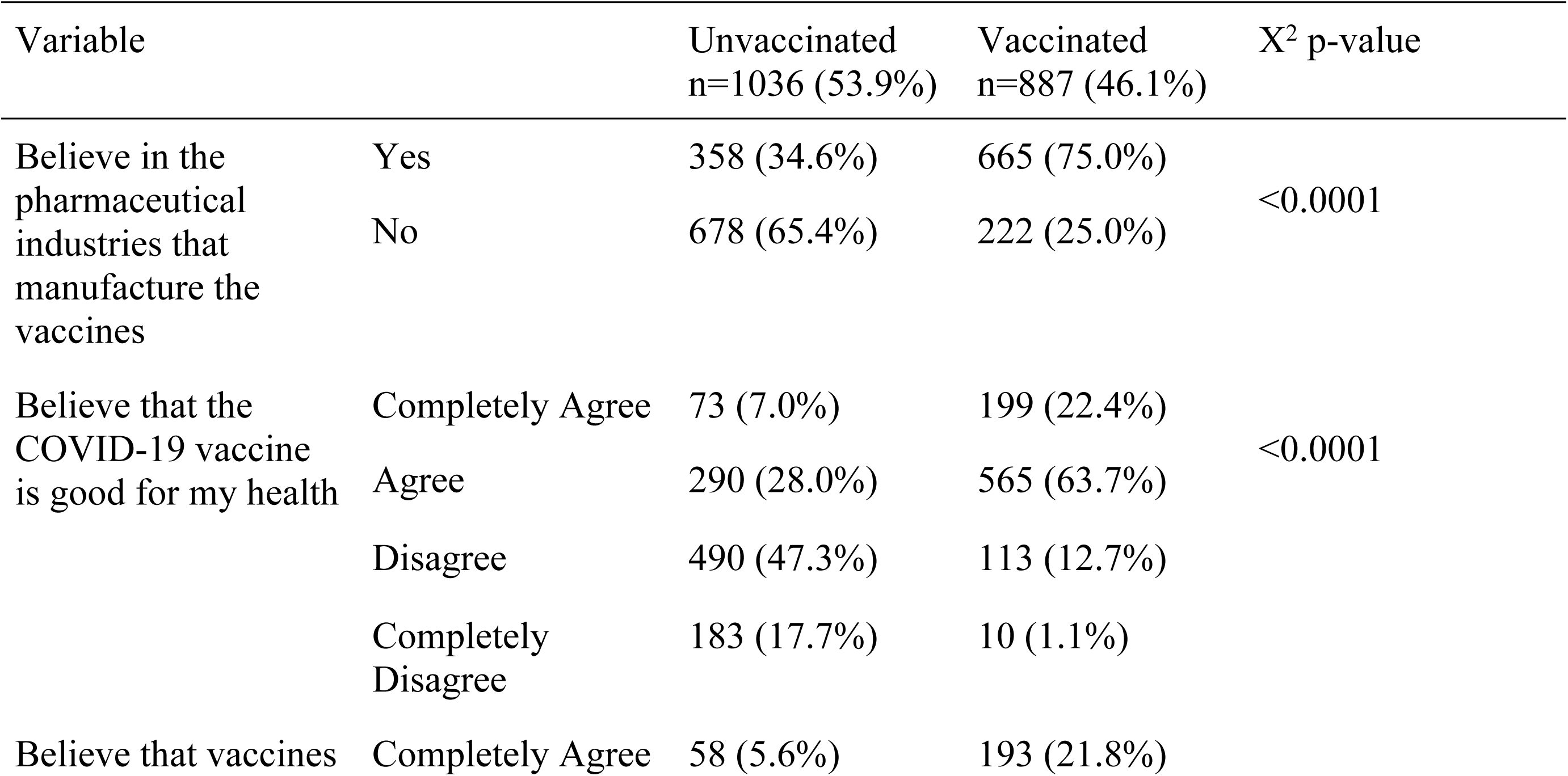
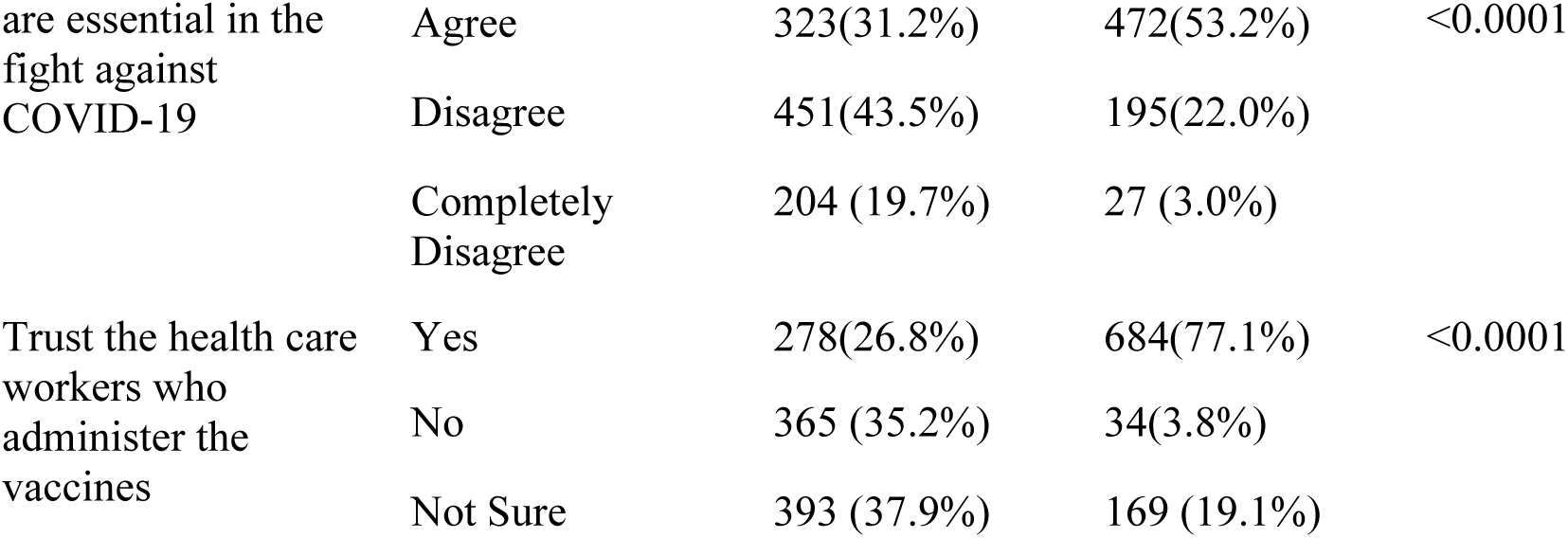
Trust in the Healthcare System and COVID-19 (N=1923)

These quantitative findings were largely supported by the qualitative results. Focus group participants shared a range of reasons for their mistrust of the health system, the Haitian government, and/or the larger global public health agenda. This is evidenced by common conspiracy theories that surfaced in different focus groups of varying vaccination status. Some participants mentioned mistrust in healthcare workers to the extent that they would not believe a COVID-19 positive test result until it had been proven by multiple tests from different locations.

Many respondents viewed the Haitian health system as ‘weak’ and expressed skepticism in the government’s messaging around COVID-19 and the vaccine. More than half (at least 6/11) of the FGDs expressed concern over nefarious motivations of the foreign governments distributing the vaccines, believing that the vaccinations were intended to cause harm to its recipients. Fear of COVID-19, misinformation, and mistrust seem to go hand-in-hand with participant’s health behaviors, causing uncertainty and skepticism towards sources that should be reliable. Health system mistrust was expressed in a number of ways from conspiracy theories to complete mistrust in the government system as a whole. This quote from one respondent represents the deep-seated mistrust expressed across many of FGD participants.

> *“I myself, if they told me that I have corona, I would say that the doctor is lying. Haitians do not stand for the [Corona]. Haiti already has its virus. Insecurity, kidnapping, they are our crown [Corona].”*

After adjusting for gender, age, education and employment status, participants who were unvaccinated for COVID were significantly more likely to have never been vaccinated for any other disease (aOR: 2.01, 95% CI: 1.32-3.05), follow anti-vaccination advice, (aOR: 2.12, 95% CI:1.26-3.55) and significantly more likely to disbelieve in vaccination effectiveness (aOR: 35.43, 95% CI: 20.73-60.58). (Table 2) Unvaccinated participants were also significantly more likely to have distrust for healthcare workers (aOR: 7.42, 95% CI:4.08-12.47) and believe that distance to the healthcare facilities was a hinderance to receiving the vaccination (aOR: 6.82, 95% CI: 2.07-22.95). Table 2.

**Table 2.**
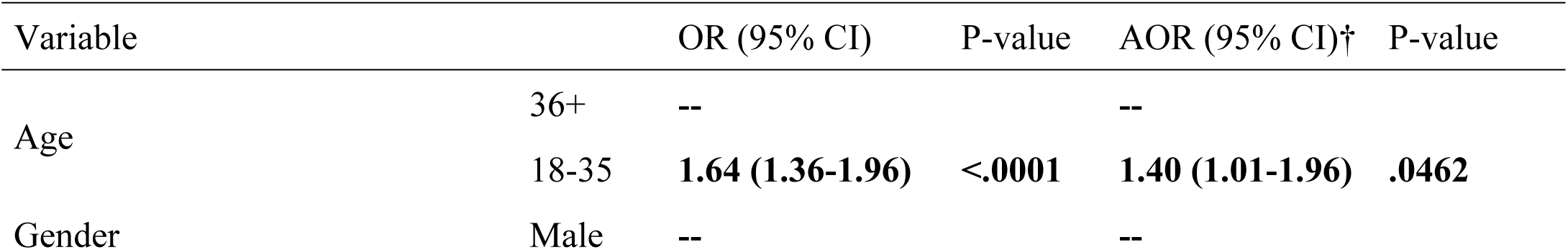
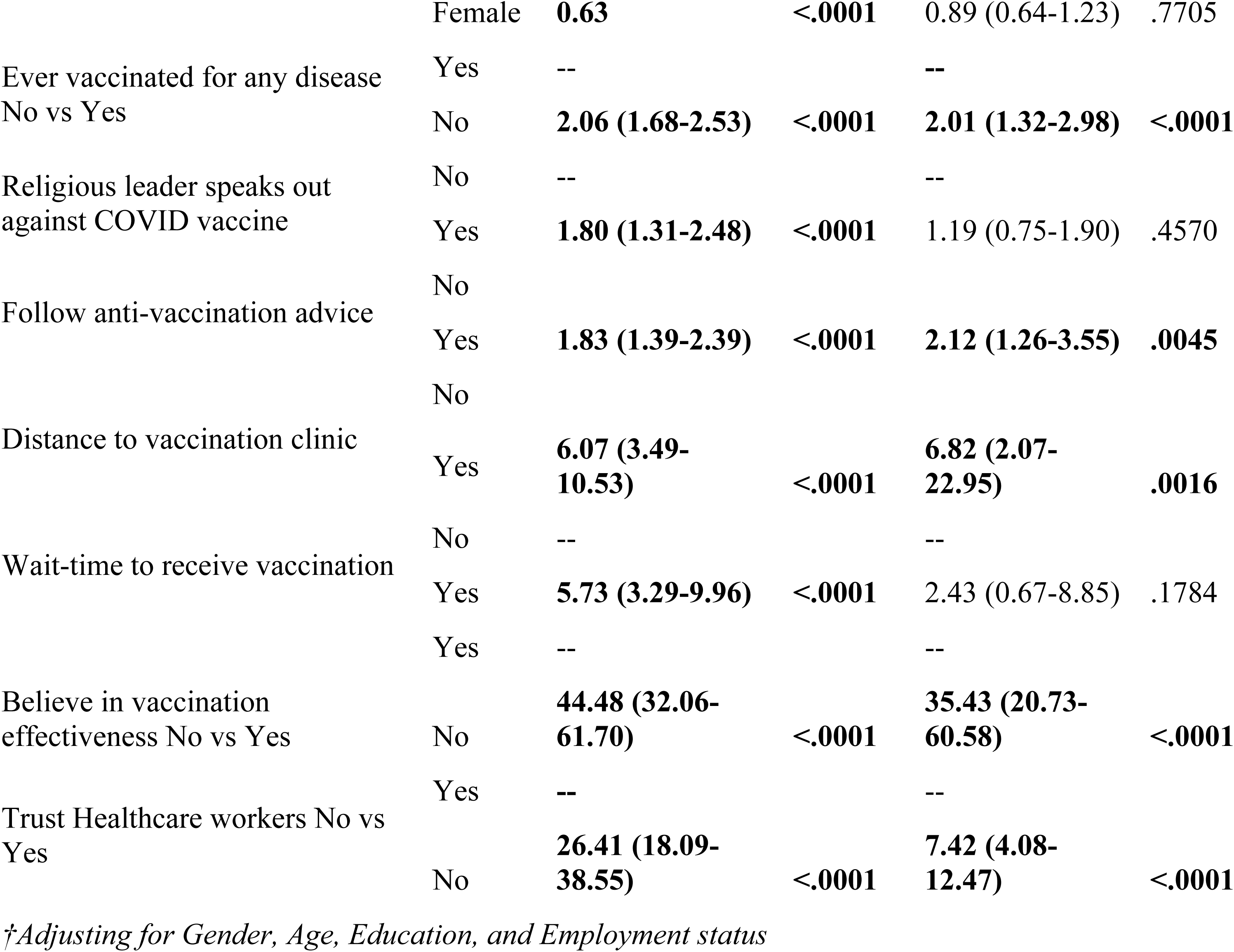
Factors Associated with COVID Vaccine Hesitancy.

## Discussion

The impact of COVID-19 was devastating across many countries. This was of particular concern for low to middle income countries struggling with poor living conditions, poor sanitation, lack of clean running water and overcrowding. Our study aimed to assess facilitators and barriers that led to vaccine hesistancy in Haiti which currently has the lowest COVID-19 vaccination rates across the Caribbean and Latin America. Left unaddressed, vaccine hesistancy can lead to a wider threat to health security jeopardizing past and future immunization gains for other preventable diseases. Our study sheds light on reasons why Haitians reported being reluctant to accepting to be vaccinated against COVID-19. Reasons also expand to knowledge and acceptance of the disease itself as well as institutional and health worker trust in the information being shared with the population. Vaccine distribution and delivery infrastructure as well as misinformation were some factors resulting in poor vaccine uptake in many low-income countries (Watson, et.al, 2022). However, even when countries gain access to vaccines the element of acceptability and trust are prevalent and must be overcome in order to decrease the emergence and spread of potentially new variants or other diseases which could have been curbed through community and population vaccinations.

A study released in 2021 found that there was a willingness to take COVID-19 vaccine in several low- and middle-income countries (Solis Arce, et. al). Since then, several studies focused primarily on low-income countries have reported a hesitancy to the COVID-19 vaccine as well as low acceptability to information surrounding COVID-19. Some of these studies found that factors associated with COVID-19 vaccine hesitancy included: perceived social norms, trust in vaccines, safety of vaccines (Davis, et. al, 2022), rapid development of vaccines, long term safety (Chukwuocha, et al, 2022), and/or distrust in pharmaceutical industry, antivaccine messages on social media (Dinga, et. al., 2021; Wouters et.,al, 2021).

Our study conducted in the northwest department of Haiti revealed similar outcomes to other studies that found that indicators such as age, gender, marital status, religion, employment status and frequency of health appointments were not shown to be predictors of receiving a COVID-19 vaccine. While our data showed that those who were not vaccinated for COVID-19 were more likely to be male (57.7%), single (57.3%), more likely to be unemployed (20.3%), hadn’t been to a medical visit in over three years (30.2%) and were likely to be less than 45 years old, when included in the adjusted model these indicators were not predictive of receiving a vaccine for Covid-19.

Our study also showed a high distrust of the healthcare workers (aOR: 7.42) and government. While Solis Arce, et. al. found that many study participants trusted in their healthcare workers and found that targeted programmatic use of healthcare workers could be used to encourage vaccination, our study found that a high distrust in healthcare workers was also connected to a distrust in government. During the implementation of our study, Haiti was experiencing political unrest coupled with a disbelief in what the government had been sharing about Covid-19 and prevention messages. This could explain the vast amount of distrust in care providers and government officials.

These findings have important implications towards programmatic rollout of mass vaccination within this population. Haiti has specific structural and political challenges that should be highlighted and our findings speak to where we should target models for solutions. For example, participants have reported low trust in healthcare workers. While our focus group discussions uncovered some reasons for distrust in healthcare workers, identifying the root of this distrust through focus group discussions for clients as well as healthcare workers would help understand the reasons behind the distrust and co-developing solutions to address the distrust. Improving patient-provider relationships, using expert patients, engaging with the community and civil society organizations and co-developing strategies with healthcare consumers allowing clients to have a stake in how their care is delivered are several opportunities to address the mistrust in healthcare providers.

There are several strategies that could be implemented to address the challenges that serve as hindrances to vaccination among this population. For example, Georgetown University with funding from the CDC Foundation implemented targeted interventions to enhance Covid-19 vaccination among the public. Working through civil society organizations, religious institutions, and community health care workers the team focused on increasing Covid-19 vaccination awareness and uptake. To address the issue of access the team supported the increase in community vaccination points working with departmental health directorates to scale up vaccinations. Further research has been focused on identifying population segments more likely to be hesistant towards vaccination with potential to move beliefs and behaviors toward vaccine confidence and uptake (Improving Vaccine Uptake, 2022). Further research to be explored could focus on a longitudinal study that follows these implementations over a period of time to assess healthcare consumer behaviors particularly around vaccinations and health seeking behaviors. In an effort to curb this “pandemic of mistrust” that is being observed across many countries, a proposed longitudinal study to review patterns of care and using targeted data to impact outcomes could prevent these identified challenges that may arise again during a future pandemic.

### Limitations

Although this study provided insight into causes of vaccine hesitancy, it was subject to a few limitations. First, it was limited to only those who were in the Western Department of Haiti between June-July 2021, so these results may not be generalizable to all Haitian residents. Approximately 1.2% of Haitian residents living in the Western department had been vaccinated by Moderna (July to November 2021) and 3% of residents had been vaccinated overall. (UEP/MSPP data (DHIS 2.0). Secondly, this was a cross-sectional study where survey responses were obtained through self-administered online questionnaires, therefore participants’ responses may have been biased. Despite these limitations, our assessment of vaccine uptake is useful for informing government and clinical entities to develop strategies to address future pandemics.

## Conclusion

Our study has identified concerns of vaccine safety, trust in healthcare providers and misinformation as key contributors to vaccine hesitancy among this population.

Future pandemic preparedness begins with addressing these factors and ensuring that the community and the healthcare workers are an integral part of this preparedness. Based on our findings, vaccine hesitancy can be minimized by working closely with community organizations and healthcare workers in order to provide accurate and evidence-based information on vaccines, their intended and unintended consequences. Transparency is important in ensuring information is shared in a trustworthy manner. Other studies have also suggested that strategies focused on providing basic messages and education on vaccines and how they work could be influential in shedding myths and poor perceptions of vaccine development and use (Burger, et. al, 2022; Jensen, et.al, 2022).

Our study findings have important implications for current and future epidemics and pandemics for Haiti’s population. With a population of over 11 million people, as of November 2022, Haiti recorded only 34,000 cases with approximately 860 deaths. Although the infection rate of Covid-19 in Haiti has been low, its Covid-19 vaccination rate is the lowest across Latin American and Caribbean countries. While we are beginning to see the global mortality curve trend downwards for Covid-19, countries with low vaccination against this virus may be at greater risk of being able to control this pandemic should it rise again. More importantly ensuring that what populations believe, how they feel and their reaction to pandemics and vaccines are a part of the strategies used to impact and influence uptake and acceptance must be front and center when safeguarding against future global health security threats. Future research is needed to examine reasons for the low infection rate in Haiti, the relationship between this low rate of infection for Covid-19 and vaccination uptake as well as the population’s thoughts on future global health security threats.

**Fig 1.**
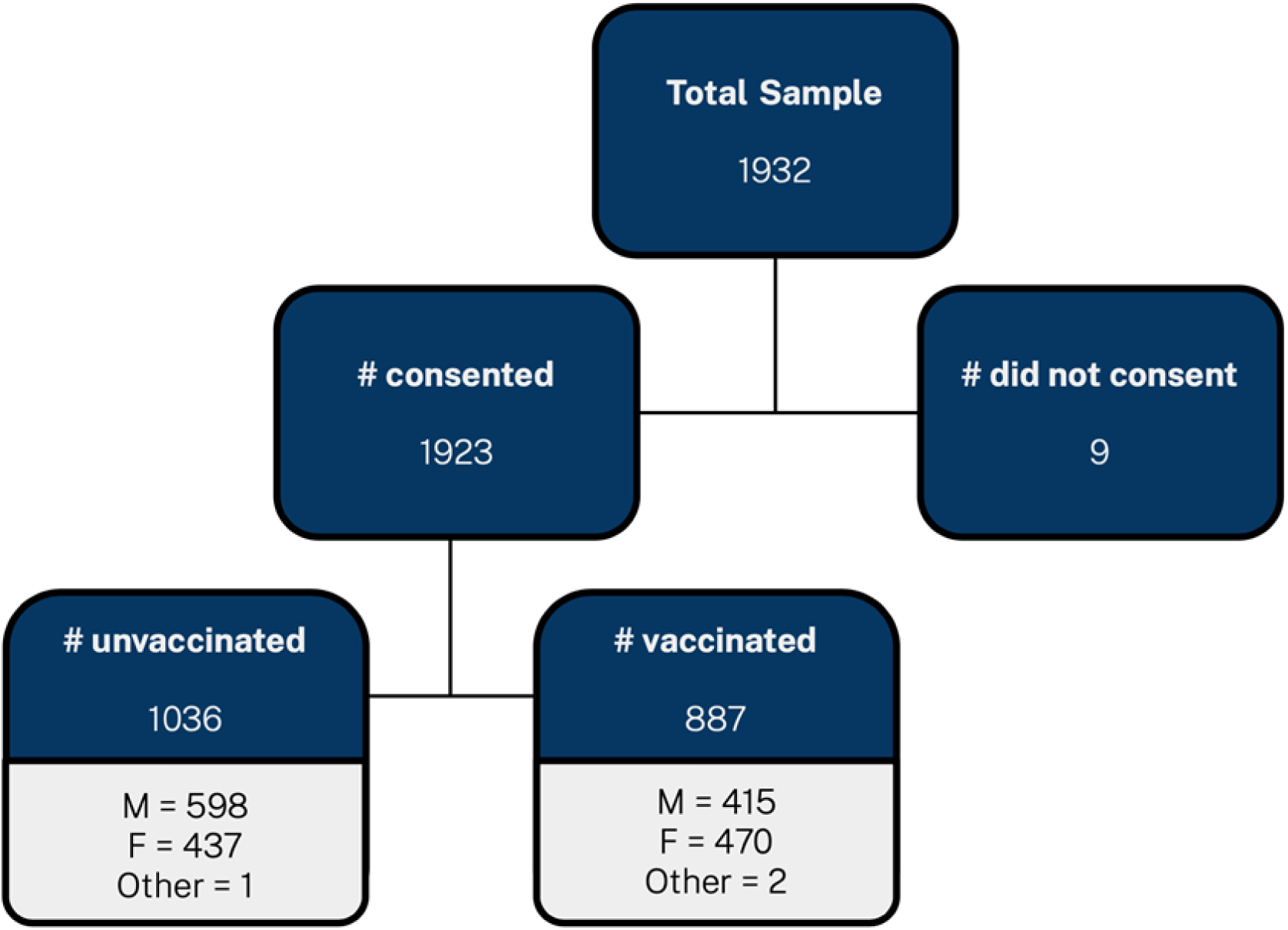
Sampling Schema.

## Data Availability

Data files will be made available according to the PLOS Global Health data policy requirements during the peer review process or before a submission can be accepted for publication.

## Acknowledgements

This manuscript is based on consultations and discussions with health care workers, community members and leaders in the community as well as technical experts and partners in support of equitable, accessible and inclusive health care. We sought and received input from the ministry of health as well as the head of the health department in the West department of Haiti. We thank the community leaders and community participants who gave voice to the issues surrounding vaccine hesistancy.

